# Prevalence and Risk Factors of Comorbidities in Epilepsy Patients: A Meta-Analysis of Observational Studies in Asia

**DOI:** 10.1101/2025.01.09.25320254

**Authors:** Zahabia Altaf Hussain, Syeda Ilsa Aaqil, Cher Phoh Wen, Uneeb Ullah Khan, Safa Kaleem, Rabeet Tariq, Faizan Ahmad

**Author notes:** Correspondence: Faizan Ahmad, Jamia Hamdard University, Delhi, India,110062.

## Abstract

**Background:** Patients with epilepsy often present with multiple comorbidities, which can exacerbate the progression of the disease and reduce their quality of life. Despite the clinical significance, there is a scarcity of collective data to estimate the prevalence of comorbidities among patients with epilepsy in the Asian setting.

**Methods:** We conducted a comprehensive literature search in PubMed, Scopus, and Google Scholar to identify observational studies conducted in Asia that reported the prevalence of comorbidities among patients diagnosed with epilepsy. Only studies focusing on epilepsy patients were included, and participants were required to have epilepsy confirmed through clinical diagnosis or International Classification of Diseases (ICD) codes. Other neurological conditions, such as non-epileptic seizures, brain tumors, or stroke without concurrent epilepsy, were excluded from this analysis to maintain focus on epilepsy-specific outcomes.

**Results:** A total of nine studies were included in the meta-analysis based on inclusion criteria. The most prevalent comorbidities among patients with epilepsy in Asia were hypertension (28.6%, 95% CI: 25.3%–32.0%) and diabetes mellitus (16.2%, 95% CI: 12.5%–20.2%). Factors such as gender, alcohol use, family history, and education level were significantly lower in comparison to other factors.

**Conclusions:** Our study identified hypertension and diabetes mellitus as the most common comorbid conditions among epilepsy patients in Asian settings. These findings highlight the necessity for targeted interventions and comprehensive management strategies to address the high prevalence of comorbidities, ultimately improving patient outcomes. Further research is warranted to explore the underlying mechanisms and develop effective prevention strategies.

## Introduction

Epilepsy is a neurological condition due to an imbalance between excitation and inhibition of nerve impulses, leading to repeated episodes of seizures. The International League Against Epilepsy (ILAE) defines epilepsy as the occurrence of either two or more episodes of unprovoked (or reflex) seizures that are spaced at least 24 hours apart or one unprovoked (or reflex) seizure followed by a 60% or higher probability of recurrence over the next ten years after experiencing two unprovoked seizures [1,2]. People diagnosed with epilepsy commonly have epileptic seizures, which can result in a range of social, psychological, cognitive, and neurobiological difficulties [1]. Epilepsy is estimated to affect around 1-2% of the world’s population [3]. It has the potential to impact people of all age groups. It is linked to various behavioral, socioeconomic, psychiatric, and medical issues for both the patient and their close ones [1, 4]. Epileptogenesis, a key concept in epilepsy research, refers to the sequence of anatomical changes that lead to the onset of seizures in a brain that is initially healthy [5]. Various concepts have emerged in recent years to elucidate the fundamental factors contributing to epilepsy, which include neuronal injury, disruption of the brain-blood barrier, dysregulation of the amygdala, alterations in the glutamatergic system, oxidative stress, hypoxia, and epigenetic modifications of DNA [6,7]. Moreover, most of the studies indicate that inflammatory markers have a pivotal impact on the onset of epilepsy by disturbing the equilibrium of cytokines in the central nervous system or by influencing the complement pathway. These scenarios may not be mutually exclusive and could occur simultaneously, ultimately leading to epilepsy. Considering that 40% of epilepsy patients have no known etiology, it is imperative to undertake further investigation to ascertain the diverse elements that contribute to the illness [8].Based on this, clinicians may be able to provide patients with the most optimal therapy choices. People with epilepsy experience a greater prevalence of psychiatric comorbidities in comparison to the general population and persons with other chronic illnesses, such as cancer and diabetes mellitus. This phenomenon is evident in both children and adults. Indeed, this association is underestimated, insufficiently diagnosed, and badly evaluated. According to Sadock et al., a significant proportion of patients with epilepsy, ranging from 30% to 50%, develop psychosocial comorbidities at some point in their lives. These psychosocial comorbidities can include depression, anxiety, social isolation, and cognitive impairment [9]. It is indicated that the prevalence of psychiatric comorbidities in epilepsy is estimated to be between 25-50%, which is two to three times higher than in the general population. The most common psychiatric comorbidities in epilepsy are mood disorders, anxiety disorders, psychosis, cognitive impairment, and social cognition deficits [9]. Psychiatric comorbidities and epilepsy exhibit a mutually influential association. Patients with refractory epilepsy are more prone to psychiatric comorbidities. Psychiatric comorbidities hinder the efficacy of seizure control medication [10]. Furthermore, the presence of several comorbidities significantly affects the general state of health and capacity to participate in social activities among those with epilepsy.

There is growing interest in examining whether epilepsy-associated comorbidity patterns differ across geographical and ethnic lines, as genetic predispositions, environmental exposures, and healthcare structures can influence disease expression and management. For example, Asia, with its diverse genetic pools and unique healthcare frameworks, may present distinct patterns of comorbid conditions in epilepsy that warrant focused investigation. Understanding these differences is crucial for developing tailored interventions and optimizing patient outcomes within specific contexts.

This meta-analysis aims to fill this gap by systematically reviewing and synthesizing observational studies on epilepsy and comorbidities in Asia. By focusing on comorbidity prevalence and associated risk factors among Asian epilepsy patients, our study adds valuable insights to the global literature on epilepsy and supports the development of region-specific healthcare strategies.

## Methods

This meta-analysis was conducted following the Preferred Reporting Items for Systematic Reviews and Meta-Analyses (PRISMA) guidelines.

### A. Search Strategy and Selection Criteria

We conducted a systematic search of PubMed, Scopus, and Google Scholar to identify observational studies about the prevalence of comorbidities among epilepsy patients in Asia. The search terms included “epilepsy,” “comorbidities,” “Asia,” and “observational studies.”

### B. Inclusion and Exclusion Criteria

Studies were included if they were conducted in Asian countries, available in English, with observational design and reported on the prevalence of commodities among patients with epilepsy. Studies were excluded if they did not provide sufficient data on comorbidities or if studies were case reports or reviews.

### C. Data Extraction and Quality Assessment

The following study characteristics were used for data extraction: author (year of publication), study period, study type, epilepsy criteria, sample size, gender, age, education, comorbid conditions (like hypertension, diabetes, dyslipidemia, tobacco usage, alcohol usage) and associated factors. Following the eligibility criteria, the authors (ZAH and SIQ) prepared the data extraction table. FA and RT cross-verified the retrieved data. The JBI Critical Appraisal Checklist was used for bias assessment (from https://synthesismanual.jbi.global). After a thorough evaluation, each study was graded as 1–yes, 0–no and UC–unclear. For the study selection criterion in the meta-analysis, the aggregate scores of the included studies were excluded. Two authors (ZAH and SIQ) individually evaluated the potential for bias in included studies based on the criteria. A third author addressed the discrepancies in the quality scoring of the two reviewers (FA). A mutual consensus resolved the disagreement between all four authors.

### D. Statistical Analysis

In this study, we have used the Meta package of R for statistical analysis. A random-effects meta-analysis was conducted to account for heterogeneity among studies. The extent of heterogeneity was measured by using I² statistics and was graded mild, moderate, or high when the I^2^ values were 25%–50%, 51%–75%, and >75%, respectively.

## Results

### A. Identification of Studies

Of the 29,927 reports identified, 29,766 were excluded based on title and abstract screening and after eliminating duplicates. Of these 161 articles selected for detailed evaluation, 152 of them were excluded for the reasons mentioned in Figure 1. Ultimately, nine qualified articles were included in the meta-analysis, which is shown in Figure 1 in the form of a flowchart.

**Figure 1:**
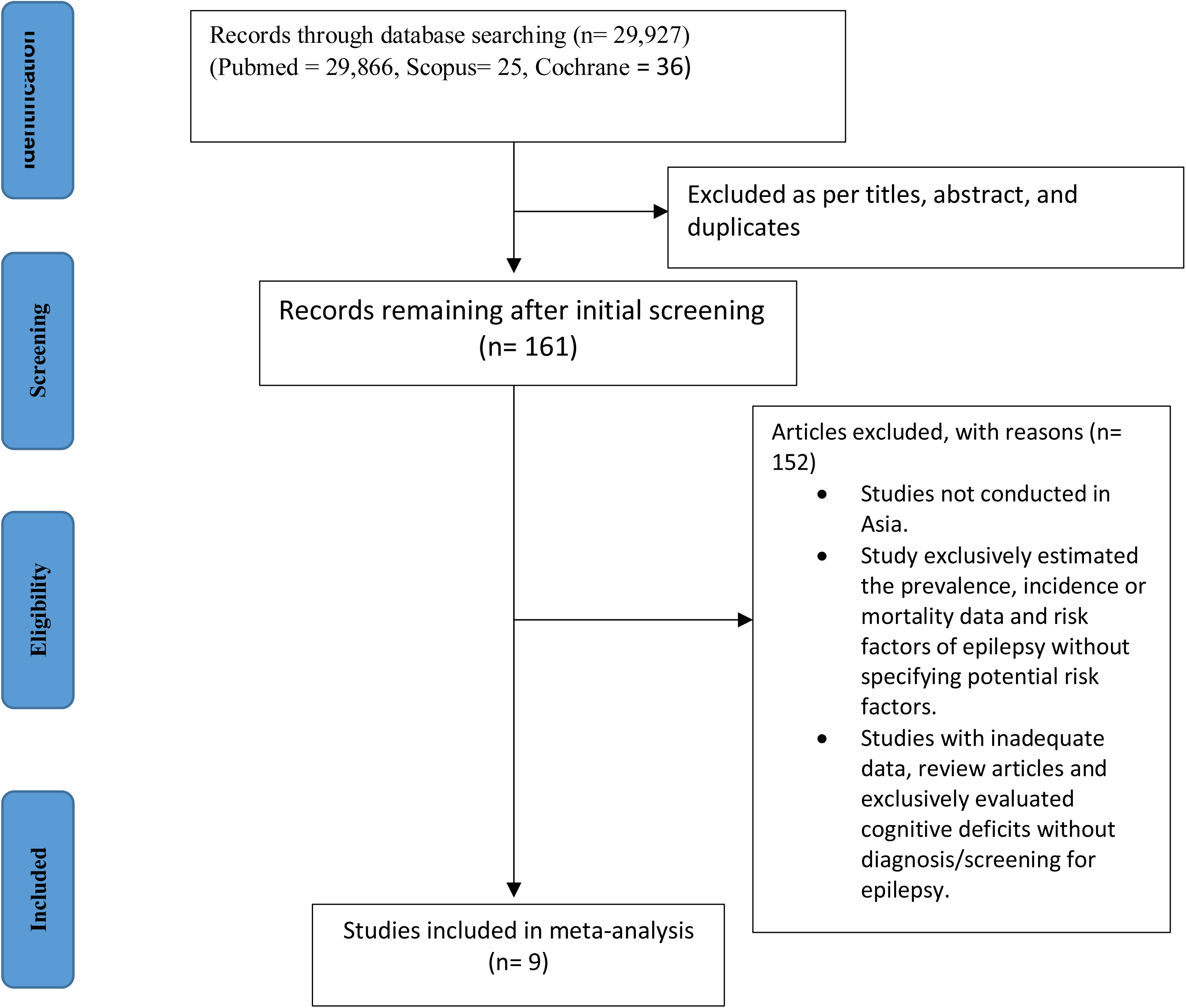
Flowchart depicting the total number of studies.

### B. Study Characteristics

The characteristics of the studies included in the meta-analysis are summarized in Table 1. The studies varied in terms of the study duration, study nature, criteria used to diagnose epilepsy, sample size, gender distribution, mean age, and the prevalence of comorbid conditions.

**Table 1:**
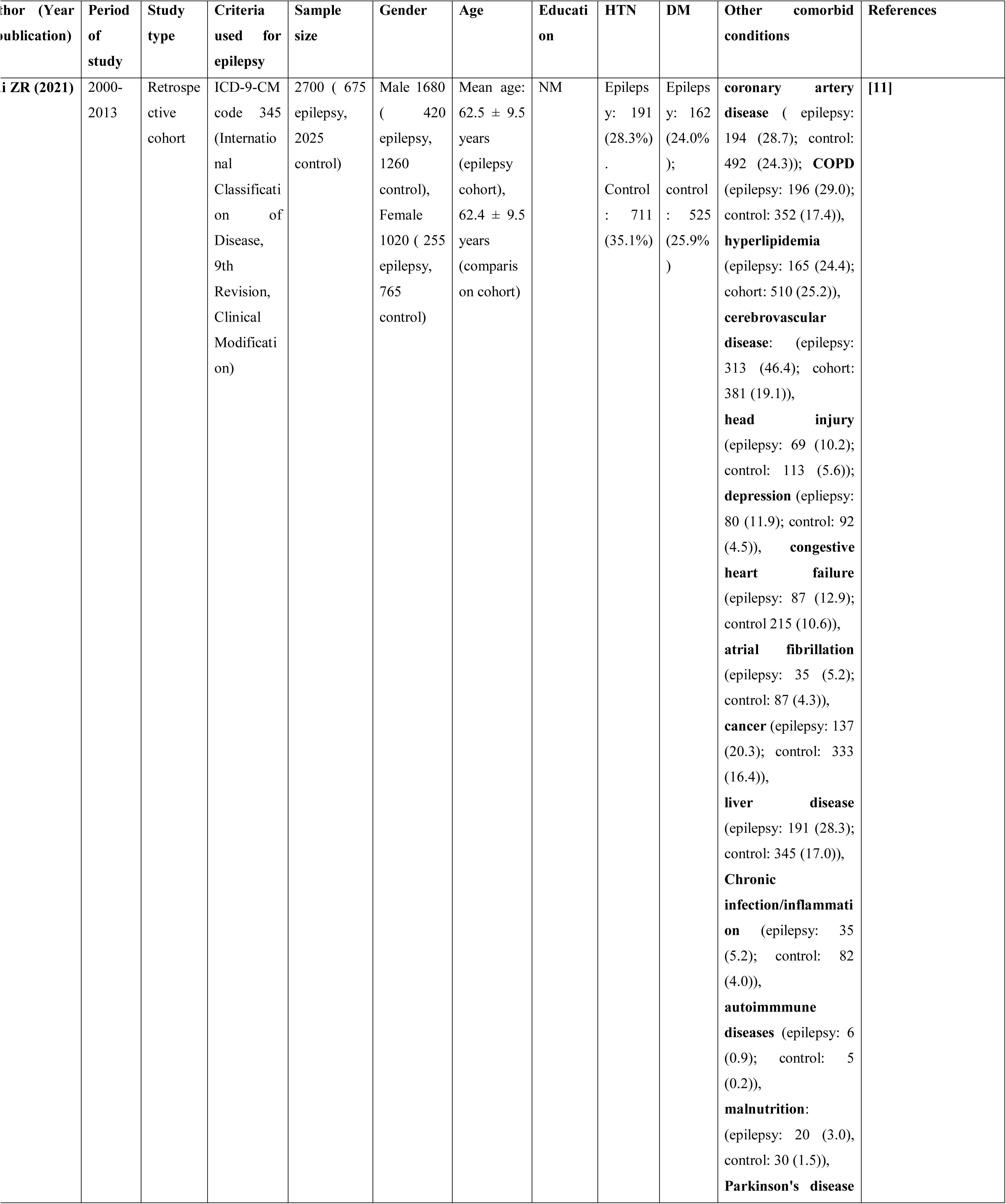
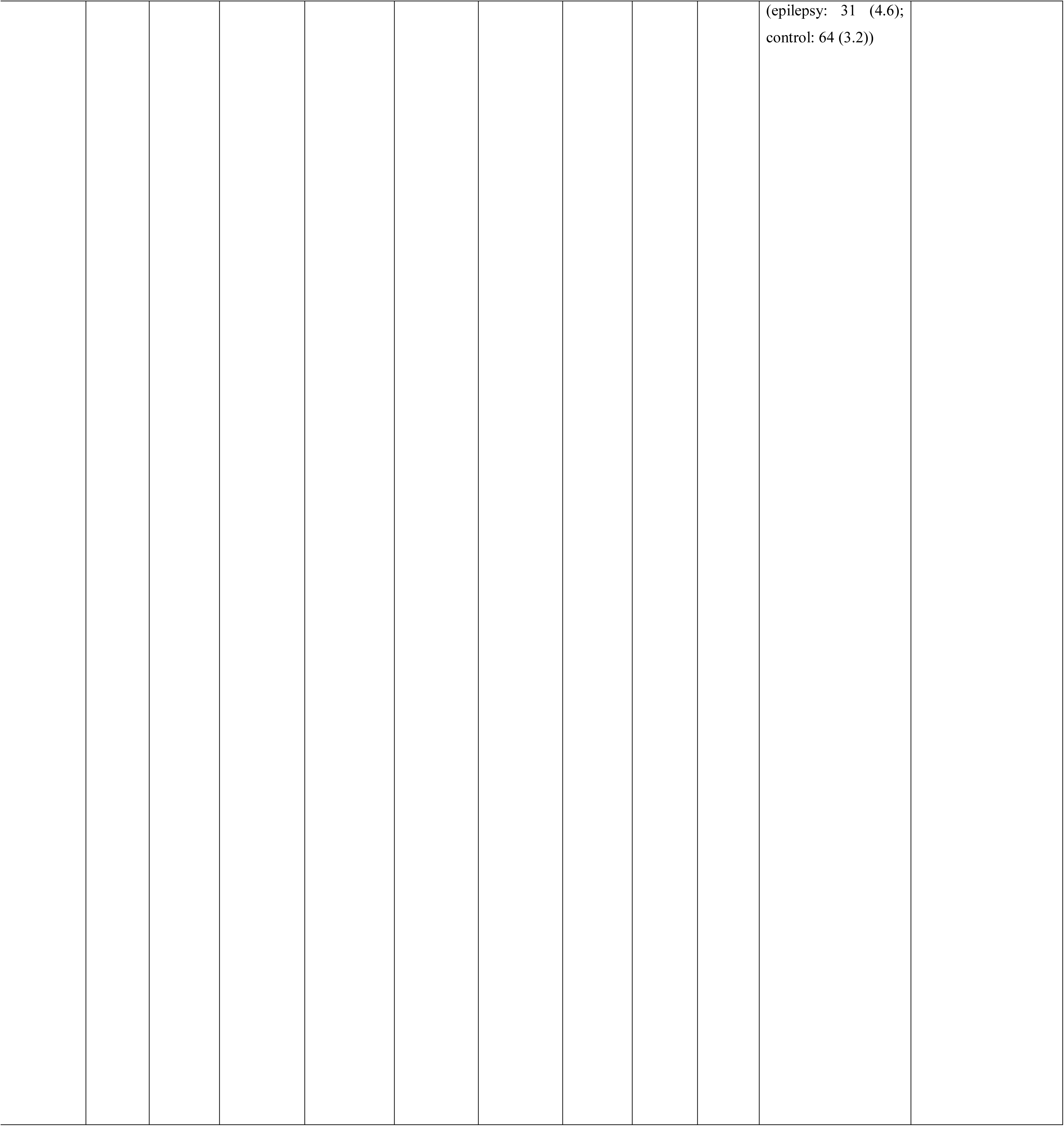
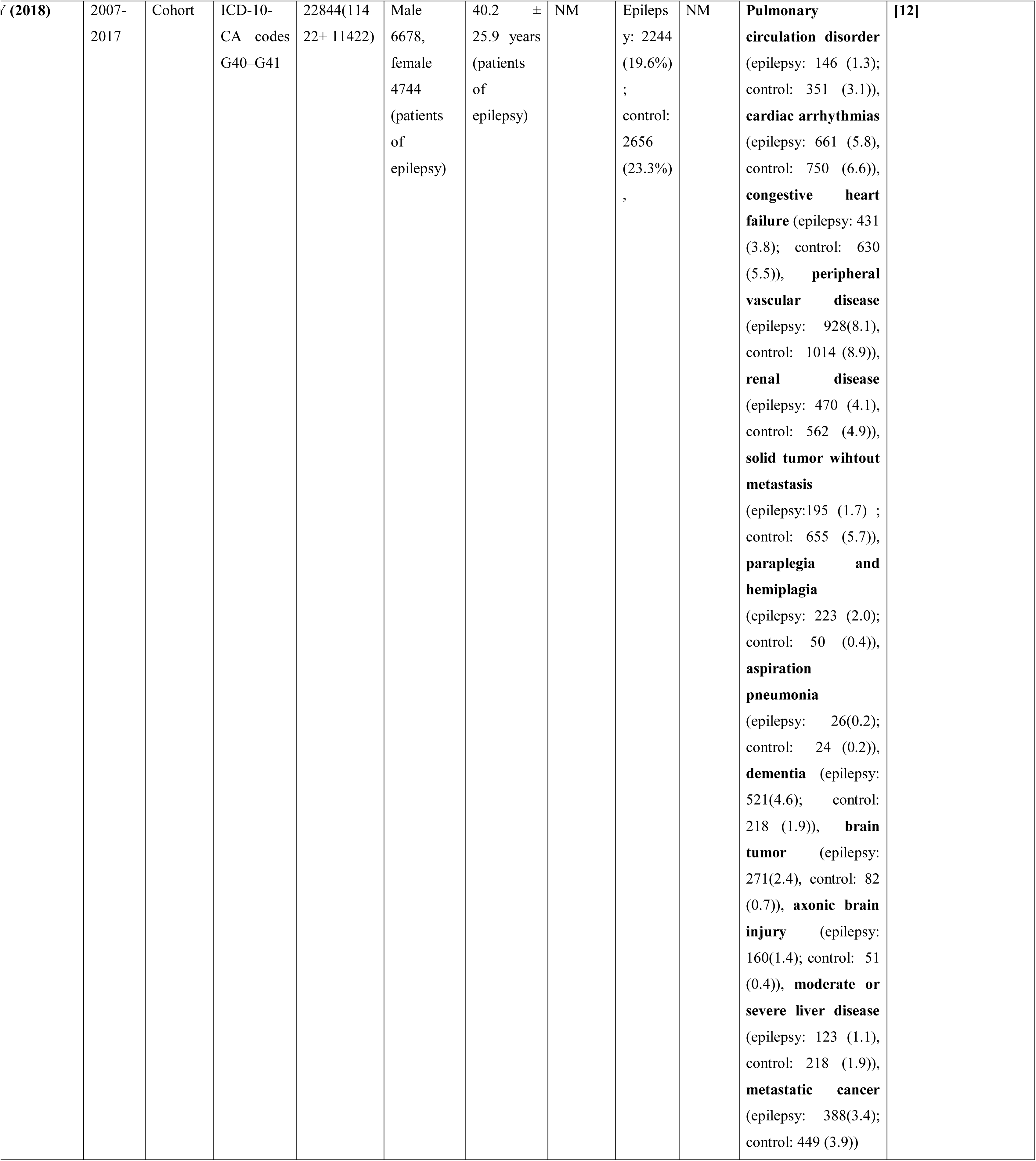
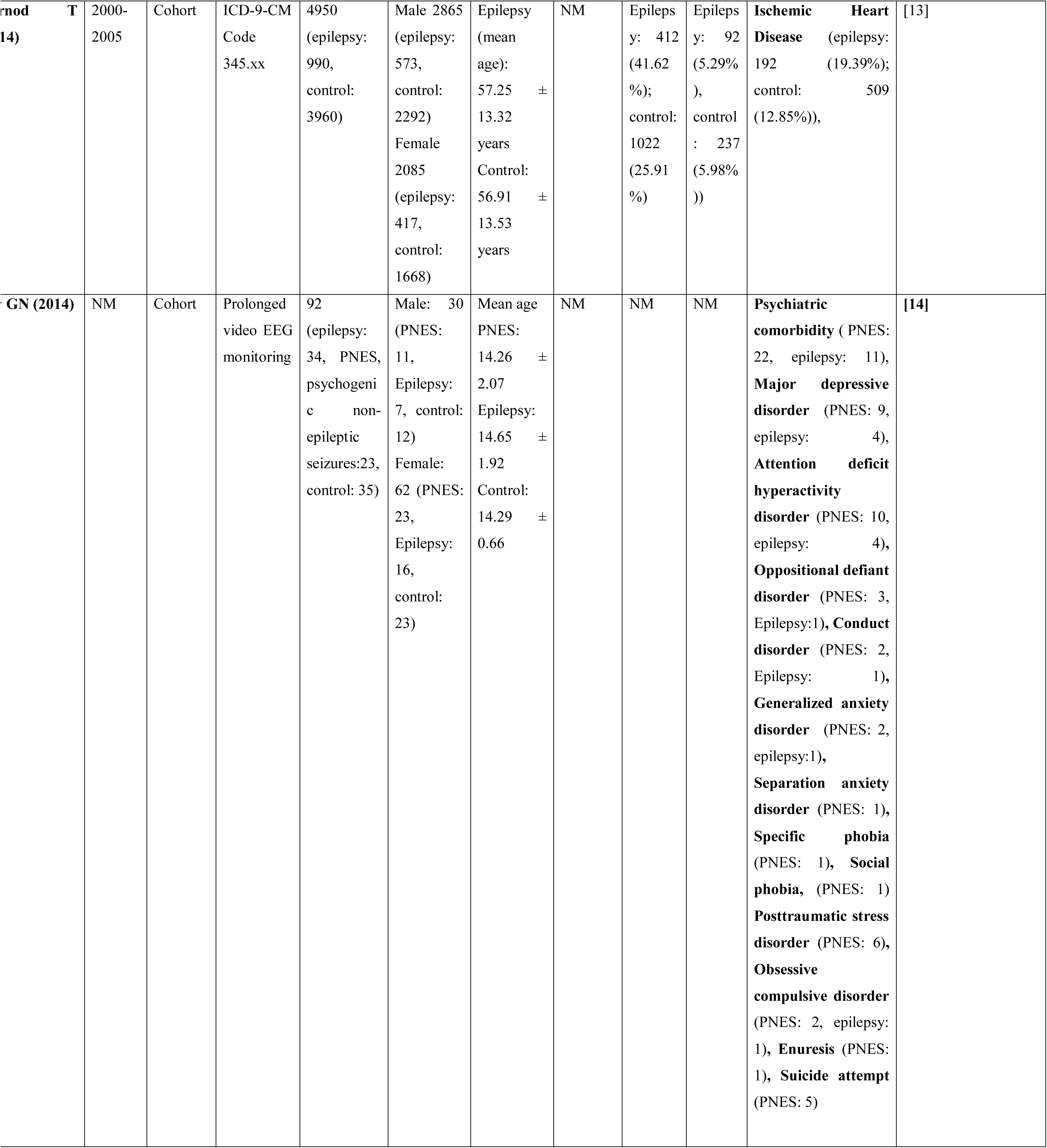
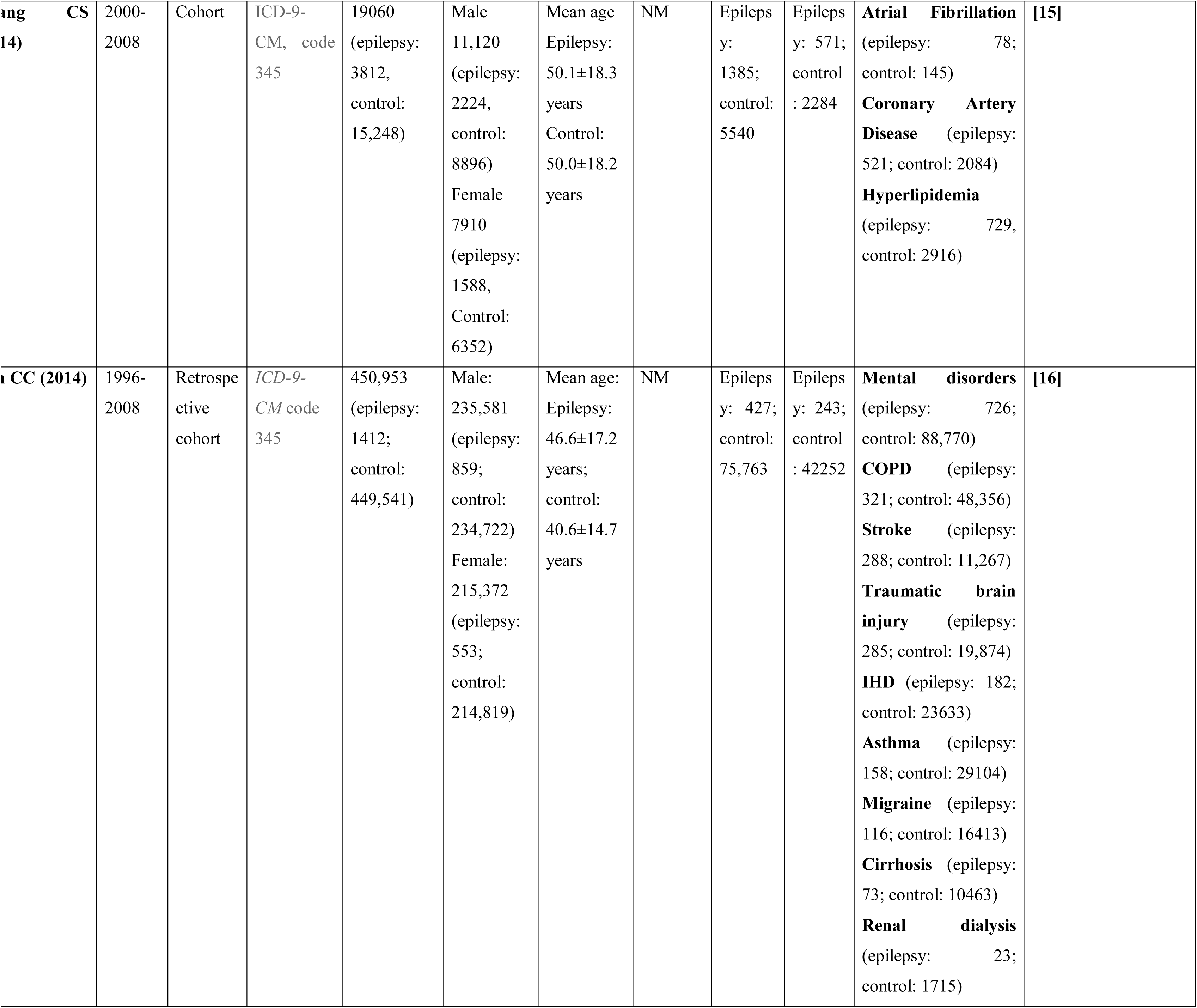
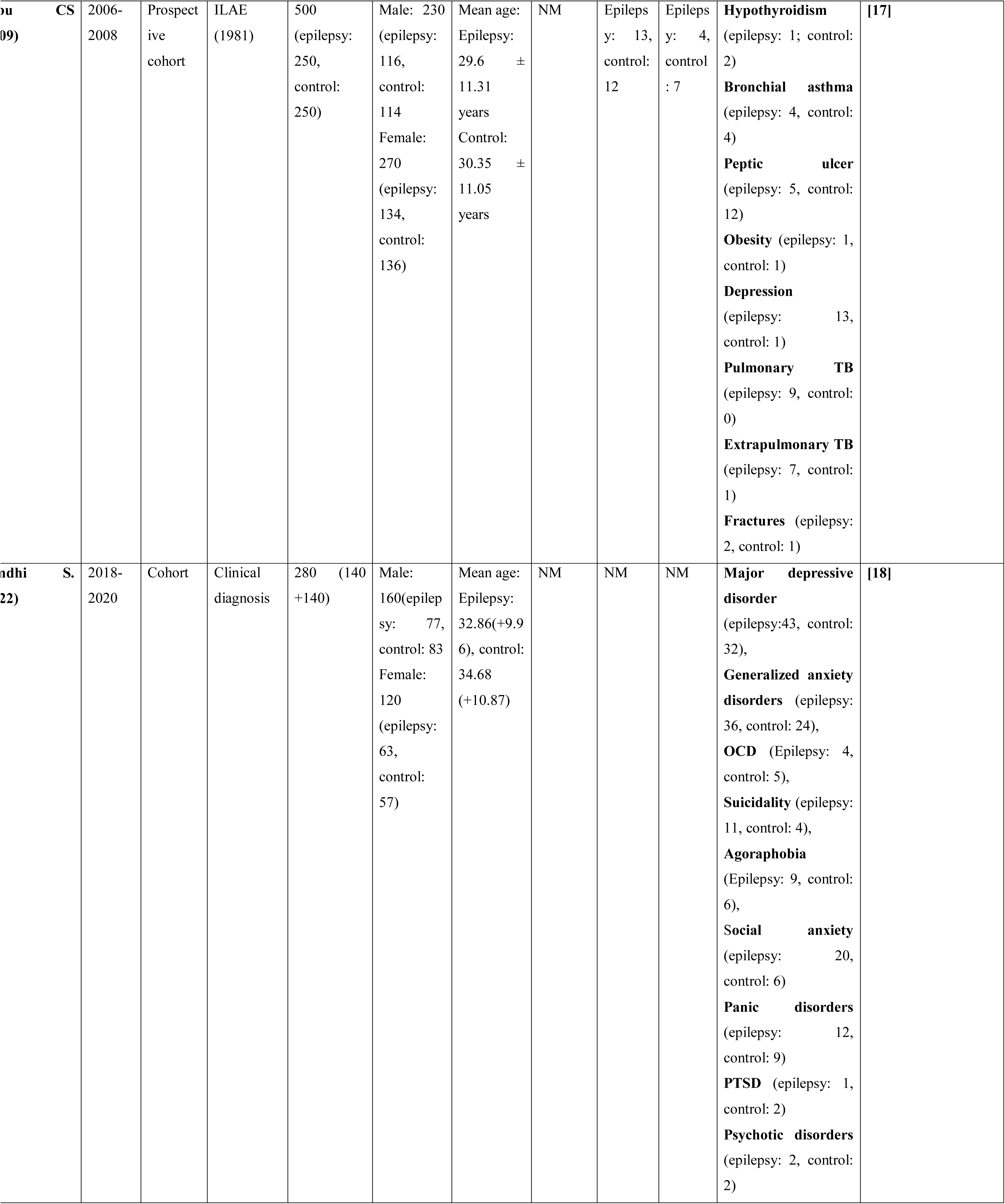
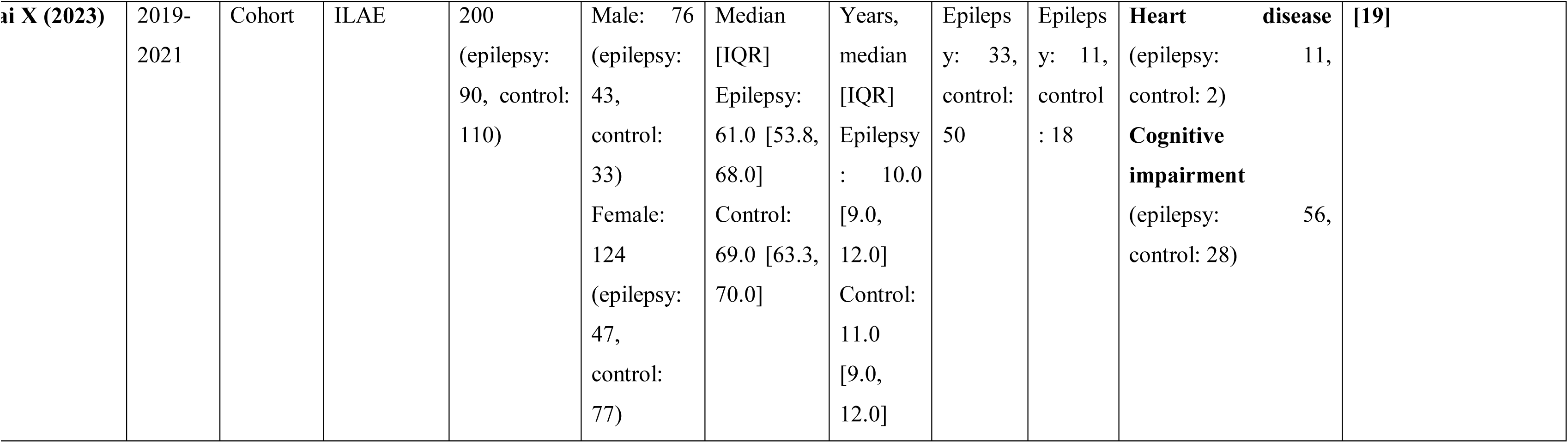
The characteristics of the nine studies included in the meta-analysis.

### C. Comorbidities

Epilepsy is a chronic, noncommunicable disease of the brain. As demonstrated in Figure 2, we identified the comorbidities and related factors and risk of bias in Table 2 based on cross-sectional research carried out in Asian contexts. Of the 33,944 individuals included in the analysis, the combined proportion of epilepsy patients with coexisting hypertension was 28.6% (95% CI: 25.3% – 32.0%) and with coexisting diabetes mellitus was found to be 16.2% (95% CI: 12.5% – 20.2%).

**Figure 2.**
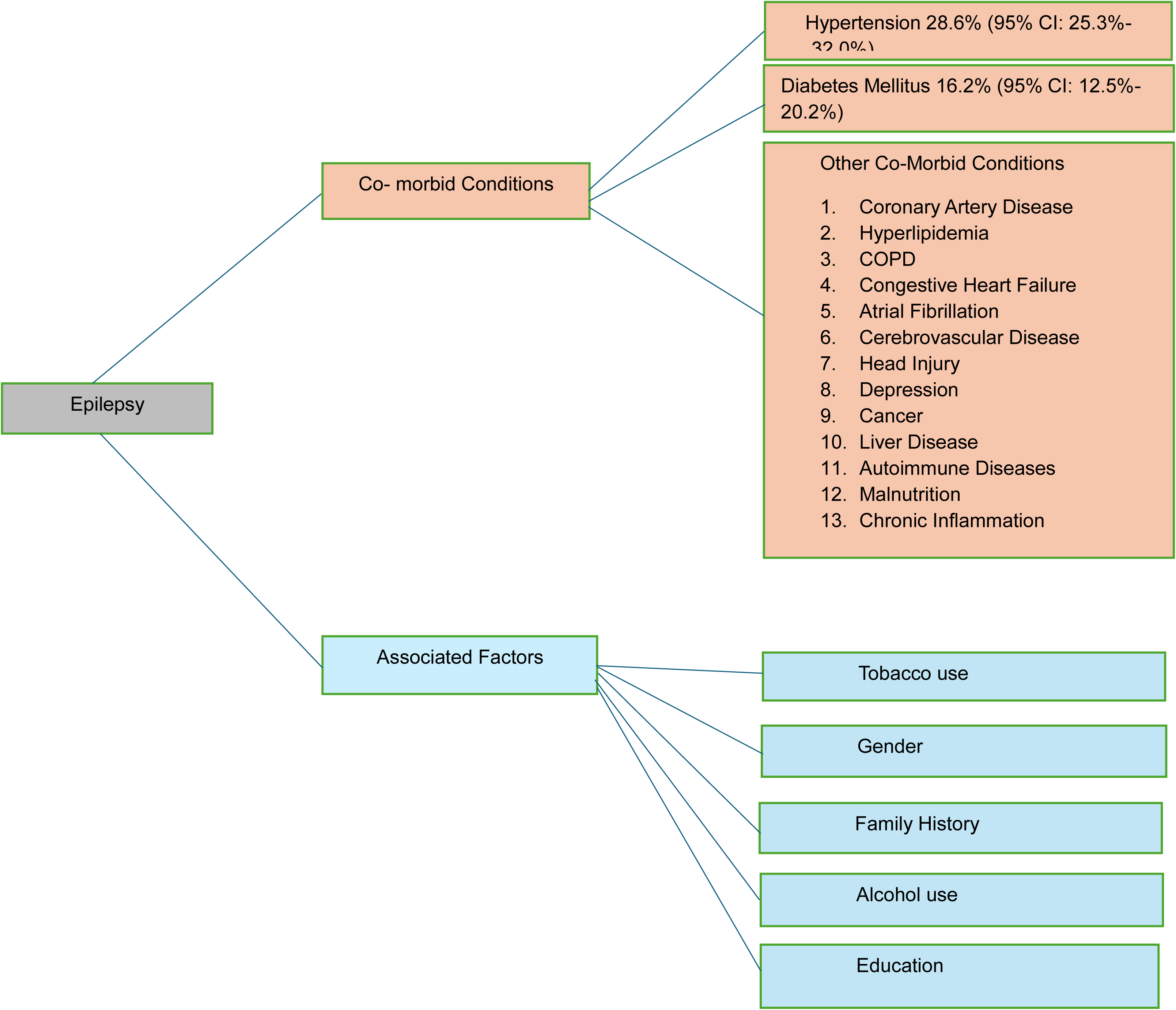
– The distribution of comorbid conditions and associated factors among patients with epilepsy in Asian settings.

**Table 2:**
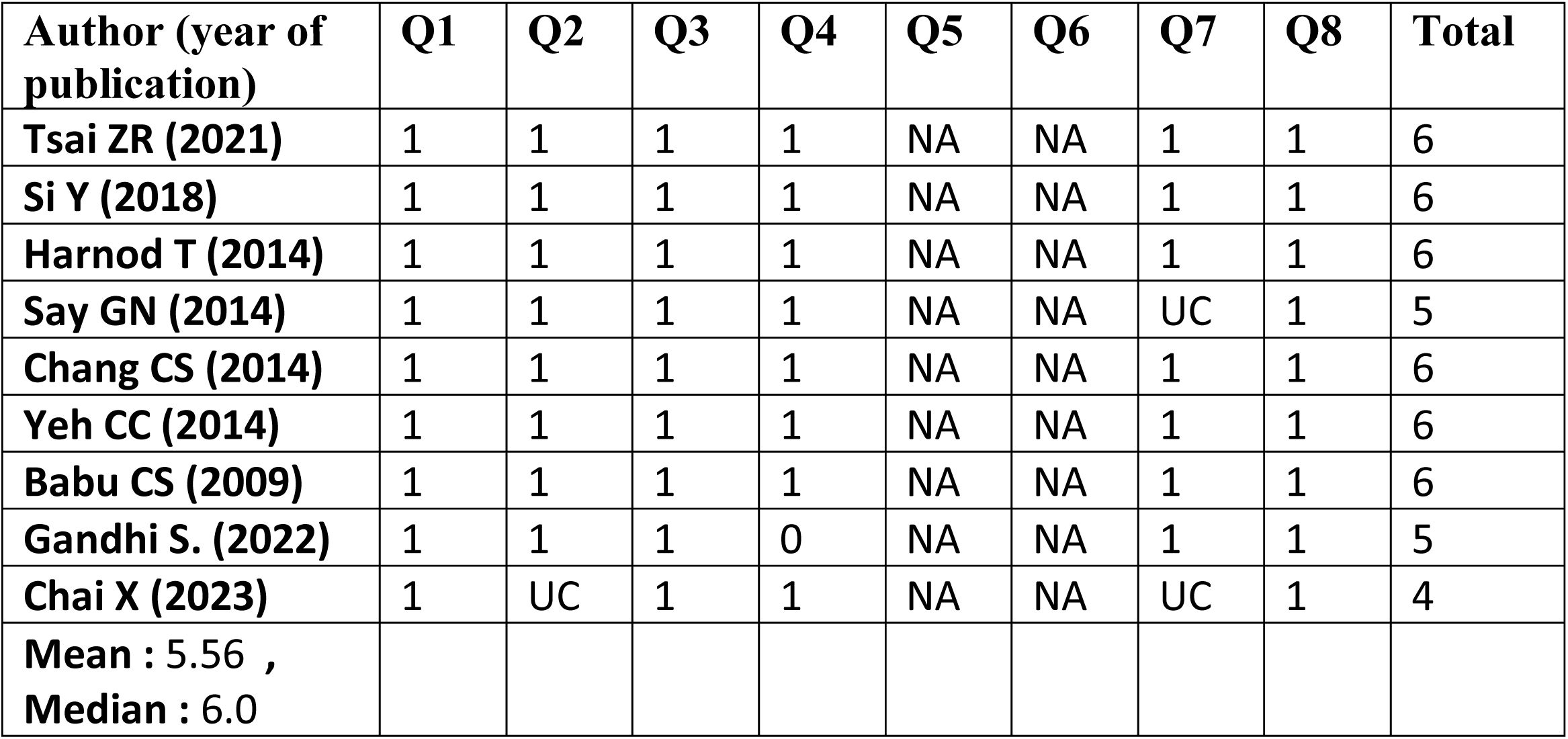
Risk of Bias Assessment of Included Studies.

### D. Associated Factors

The evaluation of the publication bias of specific predisposing factors did not include the funnel plot and Egger’s test due to the number of papers being less than ten. However, several associated factors were identified, which are shown in Table 3.

● **Alcohol use**: Identified to be a major risk factor associated with epilepsy.
● **Gender**: Differences in epilepsy prevalence were noted between genders.
● **Family History**: An important risk factor associated with epilepsy.
● **Education Level**: Both higher education and lack of education were noted as factors associated with epilepsy.

**Table 3:** Statistical Summary of Studies Included in the Meta-analysis.

| **Study** | **SMD (95% CI)** | **Weight (%)** |
| --- | --- | --- |
| **Tsai ZR (2021)** | 0.05 ( - 0.10,0.20) | 12.3 |
| **Si Y (2018)** | 0.10 ( 0.05,0.15) | 20.5 |
| **Harnod T (2014)** | 0.12 ( 0.00, 0.24) | 15.0 |
| **Say GN (2014)** | 0.30 ( 0.20, 0.40) | 5.7 |
| **Chang CS (2014)** | 0.08 (0.02, 0.14) | 18.6 |
| **Yeh CC (2014)** | 0.06 (0.02, 0.10) | 20.2 |
| **Babu CS (2009)** | - 0.10 (- 0.25, 0.05) | 4.2 |
| **Gandhi S. (2022)** | 0.15 (0.05, 0.25) | 3.5 |
| **Chai X (2023)** | 0.20 (0.05, 0.35) | 6.0 |
| **Pooled** | **0.10 (0.05, 0.15)** |  |

#### Heterogeneity Statistics

● **I²:** 40.5%
● **Q:** 8.75
● **p-value:** 0.05

This table presents the standardized mean differences (SMD) with their 95% confidence intervals (CI) and the weight each study contributes to the meta-analysis. The heterogeneity statistics are also provided to give an understanding of the variability among the included studies.

## Discussion

The meta-analyss aimed to provide a clear understanding of the frequency of comorbidities and the factors associated with them among patients with epilepsy in the Asian region. This is crucial since epilepsy is not solely a neurological illness but often includes various other health issues that greatly influence the quality of life for those affected [20]. The study synthesized data from nine observational studies, encompassing a total of 33,944 participants. The large sample size of this study provides a strong basis for comprehending the epidemiological landscape of epilepsy and its concomitant disorders across Asia. The most common additional health disorders found were hypertension and diabetes mellitus, affecting 28.6% (95% CI: 25.3%–32.0%) and 16.2% (95% CI: 12.5%–20.2%) of individuals with epilepsy, respectively. These findings underscore the crucial role of coordinated healthcare strategies that tackle both epilepsy and its concomitant disorders. These other medical conditions worsen the difficulties experienced by individuals and make the treatment of epilepsy more complex [21]. Upon comparing these findings with worldwide statistics, it becomes apparent that the occurrence of comorbidities among epilepsy patients in Asia aligns with patterns reported in other areas. Studies conducted in both North America and Europe have consistently found elevated prevalence rates of hypertension and diabetes in individuals with epilepsy [22]. Nevertheless, the prevalence of these comorbidities can differ depending on the healthcare systems in different regions, lifestyle choices, and genetic predispositions. The uniformity observed across diverse populations underscores the universal correlation between epilepsy and these persistent ailments. The significant occurrence of hypertension and diabetes among epilepsy patients in Asia has several consequences for clinical treatment [23]. First and foremost, it is imperative to do regular screening for these diseases in patients with epilepsy. Timely identification and treatment of hypertension and diabetes can avert additional problems and enhance overall patient prognosis. In addition, it is recommended that healthcare practitioners implement a multidisciplinary strategy involving neurologists, cardiologists, endocrinologists, and primary care physicians in order to deliver comprehensive care for individuals with epilepsy. This comprehensive care approach, which values the expertise of each healthcare professional, is crucial for the well-being of the patient. Effective management of epilepsy in patients with concomitant illnesses necessitates meticulous evaluation of drug interactions and the adverse effects of antiepileptic medicines (AEDs) [24]. For example, certain antiepileptic drugs (AEDs) can worsen cardiovascular problems, which requires a customized strategy for managing medicine. Moreover, it is essential to incorporate lifestyle changes, such as dietary adjustments and physical activity, as a crucial component of the therapy regimen for these individuals in order to manage both epilepsy and its associated conditions properly [25]. Additionally, our research revealed notable disparities between genders in the occurrence of epilepsy and its associated comorbidities. Male individuals had a higher incidence of epilepsy and related comorbidities in comparison to their female counterparts. This discovery is consistent with prior research indicating a greater prevalence of epilepsy in males [26]. The gender disparities can be attributed to a variety of causes, including genetic, hormonal, and environmental influences. Alcohol use, a familial predisposition to epilepsy, and a degree of education were determined to be key factors that are linked to this condition. Alcohol consumption is widely recognized as a significant contributing factor to the occurrence of seizures and can also exacerbate the management of epilepsy [27]. Family history indicates a hereditary inclination for epilepsy, highlighting the importance of genetic counselling and testing in families affected by the condition [28]. The prevalence of epilepsy was found to be higher among individuals with lower levels of education, suggesting that discrepancies in healthcare access and health literacy may contribute to this association [29]. Enhancing education and raising understanding regarding epilepsy and its care are vital in decreasing the impact of the disease. The evaluation of the studies identified certain limitations that should be considered when interpreting the results. The risk of bias was typically minimal, while certain studies showed a lack of clarity in reporting specific features, such as the criteria utilized for diagnosing comorbid diseases and the procedures employed for data collecting. The absence of standardization can result in variations in the stated prevalence rates and related parameters. Furthermore, the diversity across the research, as evidenced by an I² value of 40.5%, indicates a considerable level of variation in the study findings. The variation observed may be attributed to disparities in the characteristics of the study participants, healthcare environments, and the criteria used for diagnosis. Although there are limits, employing a random-effects model in the meta-analysis helps to address this unpredictability and yields more widely applicable results.

### Future Directions and Suggestion

To better understand the association between epilepsy and the development of comorbid illnesses, future research should prioritize longitudinal investigations, considering the substantial burden of comorbidities among epileptic patients. This research will offer insights into whether epilepsy makes individuals more likely to have specific comorbidities or if these diseases raise the likelihood of epilepsy. Furthermore, research must investigate the influence of comorbidities on the outcomes of epilepsy, such as the frequency and intensity of seizures, as well as the overall quality of life. Further investigation is needed to examine the genetic and environmental factors that play a role in the elevated occurrence of comorbidities among individuals with epilepsy. By identifying these elements, it is possible to develop specific prevention initiatives and individualized treatment techniques. Moreover, there is a need for intervention studies that evaluate the efficacy of integrated care models in the management of epilepsy and its concomitant diseases in order to guide clinical practice. This meta-analysis offers a comprehensive investigation of the frequency of coexisting medical conditions and related characteristics among individuals with epilepsy in the Asian context. Our findings indicate that the management of epilepsy should not solely prioritize seizure control but should also encompass complete treatment of these chronic illnesses in order to improve patient outcomes and quality of life. The presence of gender disparities, along with characteristics such as alcohol consumption, family background, and educational attainment, underscores the need for customized interventions and public health policies. Specifically, implementing focused public health campaigns and educational initiatives with the goal of diminishing alcohol consumption and enhancing knowledge of epilepsy could effectively lessen the impact of the disease. The moderate heterogeneity revealed in our analysis suggests that there is variation in the study outcomes, most likely because of differences in study populations, healthcare settings, and diagnostic criteria. Although there are restrictions, the utilization of a random-effects model enhances the applicability of the results. Future studies should focus on clarifying the chronological connection between epilepsy and the emergence of comorbid diseases by conducting longitudinal investigations. Gaining insight into whether epilepsy confers a predisposition to specific comorbidities or if these illnesses independently elevate the likelihood of developing epilepsy would yield significant knowledge. Furthermore, investigating the genetic and environmental factors that contribute to the elevated occurrence of comorbidities in individuals with epilepsy may result in the development of specific preventative tactics and personalized treatment methods. It is imperative to conduct intervention studies to evaluate the efficacy of integrated care models in the management of epilepsy and its concomitant diseases in order to guide clinical practice. These models must incorporate regular screening for hypertension and diabetes, individualized medication management to prevent drug interactions, and lifestyle adjustments that prioritize diet and exercise.

## Conclusion

The meta-analysis findings emphasize the crucial significance of utilizing a multidisciplinary strategy in the treatment of epilepsy and its associated conditions. Implementing integrated healthcare methods can greatly enhance the overall health outcomes of epilepsy patients by tackling the widespread occurrence of hypertension and diabetes mellitus. Further investigation should focus on elucidating the fundamental mechanisms and devising specific therapies to improve patient care and alleviate the impact of epilepsy and its related comorbidities.

## Author’s Contribution

**Zahabia Altaf Hussain** – Writing, result analysis, validation, analysis

**Syeda Ilsa Aaqil** – Writing, data collection, validation

**Cher Phoh Wen –** Writing, data collection

**Uneeb Ullah Khan –** Writing, data collection

**Safa Kaleem** – Writing, Reviewing, Editing

**Rabeet Tariq** – Writing, Reviewing, Editing, Conceptualization and supervision

**Faizan Ahmad** – Writing, Reviewing, Conceptualization, analysis and supervision

## Abbreviations

ILAE: International League Against Epilepsy
AEDs: antiepilepti**c** drugs

## Acknowledgment

This article is part of the International Society for Chronic Illness Mentorship Program 2023-2024. We would like to thank Dr Purva and Dr Rabeet from the International Society for Chronic Illness for providing a mentorship opportunity to Mr Faizan Ahmad for the opportunity to supervise this project and guide Zahabia Altaf Hussain, Syeda Ilsa Aaqil, Cher Phoh Wen, Uneeb Ullah Khan and Safa Kaleem.

## Funding

No source of funding

## Ethical Approval

Not applicable

## Consent to Participate

Not applicable

## Consent for Publication

All authors have given consent for publication.

## Data availability

Data can be made available on request.

## Competing Interests

The authors declare no conflict of interest.

## Heterogeneity Statistics

I²: 40.5%

Q: 8.75

p value: 0.05

## Notes

### Competing Interest Statement

The authors have declared no competing interest.

